# Measuring the digital divide among people with Severe Mental Ill Health using the Essential Digital Skills framework

**DOI:** 10.1101/2021.08.18.21262204

**Authors:** Spanakis Panagiotis, Wadman Ruth, Walker Lauren, Heron Paul, Mathers Alice, Baker John, Johnston Gordon, Gilbody Simon, Peckham Emily

## Abstract

**Aims:** Amidst the vast digitalisation of health and other services during the pandemic, people with no digital skills are at risk for digital exclusion. This risk might not abide by the end of the pandemic. This paper seeks to understand whether people with severe mental ill health (SMI) have the necessary digital skills to adapt to these changes and avoid digital exclusion.

**Methods:** 249 adults with SMI across England completed a survey online or offline. They provided information on their digital skills based on the Essential Digital Skills (EDS) framework, sociodemographic information, and digital access. This is the first time the EDS is benchmarked in people with SMI.

**Results:** 42.2% had no Foundation Skills and 46.2% lacked skills for daily life (lacking Foundation or Life Skills). 23.0% of those working lacked skills for professional life (lacking Foundation or Work Skills). The most commonly missing skills were handling passwords and using the device settings (Foundation Skills) and online problem solving (Skills for Life). People were interested in learning more about approximately half of the skills they did not have. People were more likely to lack Foundation Skills if they were older, not in employment, had a psychosis-spectrum disorder, or had no Internet access at home.

**Conclusion:** A significant portion of people with SMI lacked Foundation Skills in this objective and benchmarked survey. This points to a high risk for digital exclusion and the need for focused policy and tailored health sector support, to ensure people retain access to key services and develop digital skills and confidence. To our knowledge this is the first time this has been described using the Essential Digital Skills (EDS) framework. Services, including the NHS, need to be aware and mitigate the risks.

## Introduction

During the COVID-19 pandemic, restrictions have been applied to movement and social contact, to contain the spread of the virus ^1, 2^. Many daily activities (e.g. shopping, entertainment, and contacting friends and family) shifted from face-to-face to remote delivery, often via the Internet. The UK National Health Service (NHS) experienced a similar shift, both in mental ^3-5^ and general health care ^6-8^.

These changes increased the demand for digital skills and led to more people using the Internet for longer, leading to estimations that in one year the UK made five years’ worth of progress in digital engagement ^9^. However, this situation also highlighted the pre-existing digital divide (i.e. inequalities in digital access and engagement in the population) and the health inequalities that may derive from it ^10^. People lacking digital skills might experience negative outcomes in relation to health, wellbeing, and social support, as they might struggle to access health services, Government and local authority services, and support network activities that are online ^11^.

It remains unknown whether people with severe mental ill health (SMI; e.g., a diagnosis of Bipolar or Psychosis-spectrum disorder) have the digital skills to adapt to this new digitalisation of services ^12^. Finding the Internet too difficult or too complex to understand have been commonly reported among people with SMI as a barrier to use the Internet, both before ^13-15^ and during the pandemic ^16^. It is important to gain a deeper understanding of this issue, as digital exclusion in people with SMI might widen existing health and social inequalities. For example, people with SMI are at greater risk for long-term physical health conditions ^17^ and at greater need for regular health care appointment to monitor their conditions (e.g. the annual physical health check, ^18^). They might also be more likely to experience feelings of loneliness ^19^. Lack of skills to use the Internet might obstruct access to important sources of support (e.g., video-calls with health professionals or friends and family, accessing health and community related information online, and participating in online community activities). It is important to note that the risk of exclusion due to lack of skill will probably not abate at the end of the pandemic, considering that the pandemic accelerated plans for digital healthcare to become a mainstream option in the NHS ^20^

The Essential Digital Skills (EDS) framework which has been adopted by the UK Department of Education defines the skills that people would need to have to fully benefit from using the Internet and digital devices ^21^, and sets the standards for all formal digital skills training programs in the country. The framework includes three domains: Foundation Skills, Skills for Life and Skills for Work. The Foundation Skills domain concerns basic pre-requisite knowledge (e.g., knowing how to use the settings in a device, or how to connect to a secure Wi-Fi network), while the other two domains concern five skill types in everyday life and in work environments, respectively: a) Being safe, legal and confident online, b) Communicating, c) Problem solving, d) Transacting and e) Handling information and content. Although EDS is mapped with the general UK population annually through the Lloyds Bank UK Consumer Digital Index, it has never been benchmarked in people with SMI specifically.

This paper reports on the Skills and Proficiency in Digital Essential Requirements study (SPIDER) which assesses the digital skills of a sample of people with SMI amidst the COVID-19 pandemic, based on the EDS framework. The primary objectives were to understand the extent to which people with SMI have essential digital skills and how they compare to the general population, as well as to identify the areas of greatest deficit in skills and their association with key sociodemographic and health factors.

## Methods

### Design and Procedures

The SPIDER study recruited participants from a clinical cohort that participated in the Optimising Wellbeing in Self Isolation (OWLS) study. The cohort included people that were 18 years old or over and had a documented diagnosis of schizophrenia or delusional/psychotic illness (ICD 10 F20.X & F22.X or DSM equivalent) or bipolar disorder (ICD F31.X or DSM equivalent). The methods of recruitment to the OWLS study are outlined in the supplementary material. To be eligible for SPIDER, participants had to have consented during OWLS to be contacted about future surveys. Participants were recruited to SPIDER from January 2021 to March 2021.

Interested participants were provided with an information sheet (read over the phone, or send by email, text message, or post) and those consenting to participate were given the option to complete the survey online via a link, or offline (over the phone with a researcher or in a hard copy survey sent by post.) Offline options were provided to ensure inclusion of people that were not digitally engaged.

### Measures

All variables and analysis reported here have been pre-registered ^22^ in Open Science Framework (OSF).

#### Essential Digital Skills

Participants were asked whether they could complete a series of tasks related with using the Internet and digital devices, on their own if they ever need to (responses were yes or no). For those answering “no”, they were asked if they would be interested in learning more about them (yes/no). The tasks were derived from the Lloyds UK Consumer Digital Index 2020 ^23^ and the Scottish Council for Voluntary Organisations (SCVO) Essential Digital Skills Toolkit ^24^. The complete list of tasks used in SPIDER is available at https://mfr.osf.io/render?url=https://osf.io/keumd/?direct%26mode=render%26action=download%26mode=render.

Participants were considered as having or not having a given skills domain according to the EDS framework ^23^. For Foundation Skills, participants would need to report they could complete all task from a list of eights basic tasks (e.g., using a device setting to make its use easier, or connecting to a secure and safe Wi-Fi network).

For Skills for Life, we used a list of 30 tasks, grouped in five skill-types: Communicating (e.g., I can set up an email account), Handling information (e.g., I can use search engines to find information and make use of search terms to generate better results), Transacting (e.g., I can safely buy things online), Problem solving (e.g., I can use the Internet to find how to do something online), and Being safe and legal online (I am careful with what I share online as I realise that the information I put online stays there and could be accessed in the future by other people). To be considered as having Skills for Life, participants should be able to complete at least one task from each skill-type. Only participants who had Foundation Skills were assessed for Skills for Life.

For Skills for Work, we used a list of 14 tasks, grouped in the same skill-types as in Skills for Life. The difference was that the tasks in each skill-type were more relevant to a working environment (e.g., I can set up and manage an account on a professional online network/community, or I can organise, store and share work-related information on different computers, tablets or phones). To be considered as having Skills for Work participants should be able to complete at least one task in each skill-type. Only participants who had Skills for Life and were in employment were assessed for Skills for Work.

#### Digital access

In the initial OWLS survey, participants were asked if they owned a digital device (smartphones, computers, laptops or tablets - yes/no), and whether they could access the Internet from home (yes/no). In the SPIDER study we also asked whether people have used the Internet in the last 12 months (yes a lot, yes a little, or not at all) and if in the last 12 months they ever experienced not having enough data available to access the Internet as much as they would need (data poverty - yes/no).

#### Sociodemographic and health variables

Using data that were available in the initial OWLS dataset, we derived participants’ age (grouped as 18-30, 31-45, 46-65, and 66+), gender (female, male, or transgender), ethnicity (grouped as White background or Other than White), employment (grouped as being in paid employment or not) and care setting (grouped as primary or secondary mental health care). We also derived participants’ socioeconomic deprivation index according to their area of residency using their post-codes. Index scores range from 1 to 10 with higher scores indicating less deprivation and were grouped in very high deprivation (1-2), high deprivation (3-4), medium deprivation (5-6), low deprivation (7-8) and very low deprivation (9-10).

Participants’ health records were inspected for consenting participants to obtain their SMI diagnosis, which was then categorised into psychosis spectrum disorders (including schizophrenia, schizoaffective or any other psychotic disorder), bipolar disorder, or other SMI (including participants who were eligible for OWLS on the basis of a psychosis or bipolar disorder diagnosis which was later changed in their health records to something different, as for example severe depressive disorder with psychotic features). For those not providing consent to access their records or insufficient identifiable information (e.g., name and date of birth), diagnosis was coded as “not recorded”. The “not recorded” category is not reported in our pre-registered plan but was added to retain in the analysis the 48 participants falling in this group.

### Analysis

Descriptive statistics are provided for each skills domain (Foundation, Skills for Life, or Skills for Work). For participants lacking a skills domain, we present descriptive statistics for the most commonly missing skill types, and for interest in learning more about these skill types.

Association of sociodemographic and health, and digital access variables (independent variables) with having Foundation Skills were examined with a univariable binary logistic regression model before added into a hierarchical multivariable binary logistic regression. In the multivariable model all independent variables were inserted at once, with sociodemographic and health variables inserted in the first block and digital access variables in the second block. Statistical significance was set a p < .05 and analysis was conducted with IBM SPSS 26.

Some of the pre-registered analysis was not possible due to very low counts in the sample. Among people with Foundation Skills, only nine people reported having no Skills for Life (93.0%) and four working people reported having no Skills for Work (92.0%). As a result, we do not present any descriptive statistics for missing skill domains and interest in learning more among those who reported having no Skills for Work. We also did not explore which factors are associated with having Skills for Life or Work.

To test for response bias, we examined whether participants who accepted or declined our invitation to the SPIDER study differed in age, gender, ethnicity, socioeconomic deprivation, care setting and diagnosis. Differences were examined with a χ^2^ test (or the likelihood ratio if test assumptions were violated), apart for age where we used an independent samples t-test instead.

## Results

### Sample

We invited 315 adults with SMI to the SPIDER study and 249 (67.8%) participated. Those who accepted or declined the invitation did not differ in terms of any of the examined sociodemographic characteristics (Age: t(365) = -0.45, p = .650; Gender: Likelihood Ratio (2) = 4.77, p = .092; Ethnicity: χ^2^ (1) = 1.44, p = .230; Deprivation: χ^2^(4) = 6.47, p = .167; Care setting: χ^2^(1) = 0.63, p = .429; Diagnosis: χ^2^(3) = 6.07, p = .108).

Table 1 provides the sample characteristics. Participants had a mean age of 51.7 years old (range: 21-84) and the sample included 51.4% men, 46.6% women, 2% transgender, 15.6% people from other than White ethnic backgrounds, and 44.6% resided in high/very high deprivation areas. The primary diagnosis was psychosis-spectrum disorder (48.2%). The survey was completed online by 93 (37.3%) participants and over the phone or via the post by 156 (62.7%). Regarding digital access, 21.3% of the sample were non-users of the Internet, 12.4% did not own a digital device, 15.3% had no access to the Internet at home, and 11.2% had experienced data poverty.

**Table 1.** Sample characteristics (N = 249)

| Variable (valid N) | N (%) |
| --- | --- |
| **Age (249)** |  |
| 18-30 | 28 (11.2) |
| 31-45 | 64 (25.7) |
| 46-65 | 100 (40.2) |
| 66+ | 57 (22.9) |
| **Gender (249)** |  |
| Female | 116 (46.6) |
| Male | 128 (51.4) |
| Transgender | 5 (2.0) |
| **Ethnicity (249)** |  |
| Asian | 14 (5.6) |
| Black | 4 (1.6) |
| Mixed | 11 (4.4) |
| White British | 200 (80.3) |
| White (other) | 10 (4.0) |
| Other | 10 (4.0) |
| **Socioeconomic Deprivation (241)** |  |
| Very high | 60 (24.1) |
| High | 51 (20.5) |
| Medium | 49 (19.7) |
| Low | 43 (17.3) |
| Very low | 38 (15.3) |
| **In paid employed (248)** |  |
| Yes | 61 (24.5) |
| No | 187 (75.1) |
| **Diagnosis (249)** |  |
| Bipolar disorder | 83 (33.3) |
| Psychosis spectrum disorder | 120 (48.2) |
| Other SMI | 16 (6.4) |
| Not recorded | 30 (12.0) |
| **Care setting (247)** |  |
| Primary care | 95 (38.2) |
| Secondary care | 152 (61.0) |
| **Using the Internet** |  |
| A lot | 137 (55.0) |
| Just a little | 58 (23.3) |
| Not at all | 53 (21.3) |
| **Owning any digital device (245)** |  |
| Yes | 214 (85.9) |
| No | 31 (12.4) |
| **Access to the Internet at home (246)** |  |
| Yes | 208 (83.5) |
| No | 38 (15.3) |
| **Ever felt not having enough data? (248)** |  |
| Yes | 28 (11.2) |
| No | 177 (71.1) |
| N/A (Not using the Internet) | 43 (17.3) |

### Essential Digital Skills

Table 2 presents the EDS framework descriptive results. Out of the total sample (N = 249), 105 participants (42.2%) reported no Foundation Skills. They were unable to complete on average 3.64 (±2.50) tasks out of eight in total but were interested in learning more about on average 2.22 (±2.80) of them. The tasks that people who reported no Foundation Skills were most often unable to perform were updating and changing passwords (68.6%) and using the device settings to improve its usability (e.g. change font sizes, screen brightness, screen contrasts, etc.) (61.9%). The least frequent skill deficit was not knowing how to turn on a device (17.1%) (Figure 1).

**Table 2.** Essential Digital Skills Framework.

|  | N (%) |
| --- | --- |
| **Skills Domains** |  |
| **Foundation Skills <sup>a</sup>** |  |
| Yes | 142 (57.0) |
| No | 105 (42.2) |
| **Skills for Life <sup>b</sup>** |  |
| Yes | 132 (93.0) |
| No | 9 (6.3) |
| **Skills for Work <sup>c</sup>** |  |
| Yes | 46 (92.0) |
| No | 4 (8.0) |
| **Overall** |  |
| **Essential skills for daily life <sup>a</sup>** |  |
| Yes (having Foundation AND Life Skills) | 132 (53.4) |
| No (Lacking Foundation OR Life Skills) | 114 (46.2) |
| **Essential skills for professional life <sup>d</sup>** |  |
| Yes (Having Foundation AND Work Skills) | 46 (75.4) |
| No (Lacking Foundation OR Work Skills) | 14 (23.0) |

a. Out of total sample N = 249. b. Out of those having Foundation Skills, n = 142. c. Out of those being in paid employment and having Skills for Life, n = 50. d. Out of those being in paid employment, n = 61.

**Figure 1.**
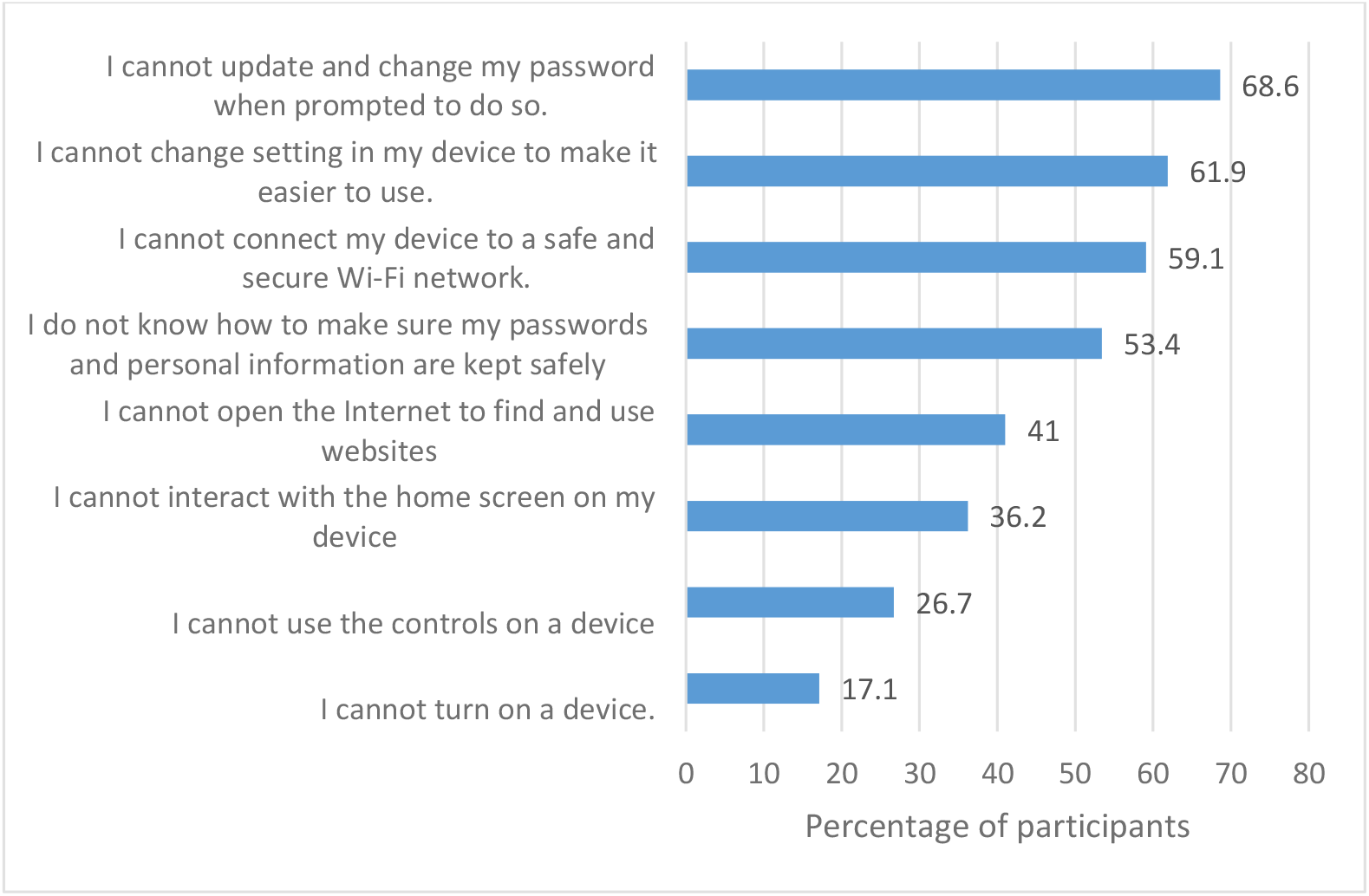
Prevalence of skills deficiency among participants with no Foundation Skills (N = 105)

Overall, 114 participants from the total sample (45.7%) did not have the essential digital skills for daily life, lacking either Foundation Skills or Skills for Life. Among the working population in our sample (N = 61), 14 participants (23.0%) did not have the essential digital skills for professional life, lacking either Foundation Skills, Skills for Life, or Skills for Work.

### Sociodemographic associations with Foundation Skills

A multivariable binary logistic regression model (Table 3) demonstrated that reporting having Foundation Skills was eight and four times more likely among the 18-30 and 31-45 age groups respectively (compared to 66+), three times more likely among those in paid employment, and four times more likely in those with a bipolar disorder diagnosis (compared to psychosis-spectrum). It was also found that participants with no Internet access at home were seven times less likely to report they had Foundation Skills.

**Table 3.** Association of having Foundation Skills with sociodemographic and digital access variables.

| Variables (Valid N) | Have Foundation Skills | Univariable Model |  | Multivariable Model (N = 229) |  |
| --- | --- | --- | --- | --- | --- |
|  | N (%) | OR | 95% CI | AdjOR | 95% CI |
| **Age (247)** |  |  |  |  |  |
| 18-30 | 24 (85.7%) | 10.29** | 3.14 – 33.72 | 7.56* | 1.75 – 32.66 |
| 31-45 | 45 (70.3%) | 4.06** | 1.90 – 8.68 | 3.60* | 1.22 – 10.57 |
| 46-65 | 52 (53.1%) | 1.94 | 0.99 – 3.78 | 1.93 | 0.75 – 4.97 |
| 66+ | 21 (36.8%) | 1 |  | 1 |  |
| **Gender (242)** |  |  |  |  |  |
| Male | 74 (58.3%) | 1.07 | 0.65 – 1.79 | 1.78 | 0.89 – 3.53 |
| Female | 65 (56.5%) | 1 |  | 1 |  |
| **Ethnicity (247)** |  |  |  |  |  |
| Other than White | 22 (56.4%) | 1.05 | 0.53 – 2.10 | 1.05 | 0.43 – 2.60 |
| White | 120 (57.7%) | 1 |  | 1 |  |
| **Socioeconomic deprivation (239)** |  |  |  |  |  |
| Very high | 29 (49.2%) | 0.63 | 0.28 – 1.44 | 0.65 | 0.22 – 1.97 |
| High | 26 (52.0%) | 0.71 | 0.30 – 1.66 | 0.44 | 0.14 – 1.36 |
| Medium | 34 (69.4%) | 1.48 | 0.61 – 3.60 | 1.37 | 0.44 – 4.23 |
| Low | 24 (55.8%) | 0.82 | 0.34 – 2.00 | 0.56 | 0.18 – 1.73 |
| Very Low | 23 (60.5%) | 1 |  | 1 |  |
| **In paid employment (247)** |  |  |  |  |  |
| Yes | 50 (83.3%) | 5.16** | 2.47 – 10.79 | 2.67* | 1.07 – 6.66 |
| No | 92 (49.2%) | 1 |  | 1 |  |
| **Diagnosis (247)** |  |  |  |  |  |
| Not recorded | 19 (63.3%) | 1.94 | 0.85 – 4.44 | 1.40 | 0.48 – 4.07 |
| Other SMI | 9 (60.0%) | 1.69 | 0.57 – 5.04 | 1.43 | 0.40 – 5.17 |
| Bipolar | 58 (69.9%) | 2.61* | 1.45 – 4.71 | 3.67* | 1.56 – 8.64 |
| Psychosis spectrum | 56 (47.1%) | 1 |  | 1 |  |
| **Care setting (245)** |  |  |  |  |  |
| Secondary care | 86 (57%) | 0.94 | 0.56 – 1.58 | 1.15 | 0.55 – 2.38 |
| Primary care | 55 (58.5%) | 1 |  | 1 |  |
| **Device ownership (243)** |  |  |  |  |  |
| No | 2 (6.5%) | 0.04** | 0.01 – 0.16 | 0.26 | 0.04 – 1.70 |
| Yes | 138 (65.1) | 1 |  | 1 |  |
| **Internet access at home (244)** |  |  |  |  |  |
| No | 3 (7.9%) | 0.04** | 0.01 – 0.15 | 0.14* | 0.03 – 0.70 |
| Yes | 137 (66.5%) | 1 |  | 1 |  |

\* p < .05, \*\* p < .001

## Discussion

We assessed digital skills in a sample of people with SMI using the Essential Digital Skills framework. To our knowledge this is the first time this benchmark measure of knowledge and skills has been applied in a sample of people who use mental health services. It is concerning that 42% (n = 105) had no Foundation Skills, lacking pre-requisite knowledge to interact with digital technologies and benefit from its use. The most problematic areas were handling passwords and using device setting to improve usability. Older people, those not currently in work, those with a psychosis-spectrum disorder, and those with no Internet access at home were at greater risk for lacking Foundation Skills. More positively, among people who had Foundation skills, almost everyone had Skills for Life and Skills for Work.

Our results reveal a digital divide between our SMI sample and the general population. People with SMI in this study where twice as likely to experience a deficit in either Foundation or Life Skills (n = 114, 46.2%) compared to the general population (22% ^23^). During the pandemic, this might contribute to inequalities of access to digital services, including but not limited to healthcare services, due to lack of familiarity or confidence with technology ^25^. More worryingly, these inequalities might remain beyond the pandemic ^26^, given that transition to digital healthcare might become a more mainstream option ^20, 27^. Combined with the fact that most people in our study owned a digital device (n = 214, 85.9%), this implies that access to digital devices on its own many not mitigate for digital exclusion and that many people may use their devices only for very basic things. This suggests that digital skills training programs are needed, that will be tailored to the needs of people with SMI, to help close this gap (see for example Recovery Colleges at https://imroc.org/resources/1-recovery-colleges/).

It was interesting that fewer of the working people in this study experienced a deficit in skills necessary for work, lacking either Foundation or Work Skills (n = 14, 23%), compared to the general population (52% ^23^). This could be a sampling issue in our study and might not be representative of the total SMI population. However, it may also be explained by the fact that employment is less common in people with SMI, ^28-30^, as shown by the 75.1% (n = 187) rate of unemployment in this study, and has been associated with younger age and higher educational attainment ^29, 31, 32^, as well as less severe SMI symptoms ^29, 32^. All of these might help people to acquire good digital skills and this may be directly linked with this study’s finding that employed participants were more likely to have Foundation Skills compared to those not in employment. Future studies should explore Skills for Work among people with SMI who are unemployed, as an indicative factor of their employment prospects and potential learning needs. Researchers should also seek to understand the contribution of digital skills deficits in the high unemployment rates in people with SMI (e.g., most jobs advertised online), especially in relation to other employment barriers related to SMI (e.g. ill health or stigma).

Looking further into skills deficits, we found that people with no Foundation Skills struggled the most with handling passwords and using device settings. As such, important as it is to providing people with digital devices to tackle access issues (as happened a lot during the pandemic; ^33^), this should be combined with training and support for using these devices. Moreover, creating and maintaining a password is imperative to access several online services (e.g. GP services, streaming platforms, online banking and Government online services, to name a few), so that deficits in that area may adversely affect not only people’s confidence in using these services but also to the security of their information. As these skills seem to be often missing in the general population as well ^23^, it is important for future studies to explore the specific barriers the SMI population might face, and how to address these from a person-centred design perspective. Among the few people who had Foundation Skills but no Skills for Life, the greatest area of deficit was Problem Solving skills. Evidence suggests that lack of problem solving skills in general, not just digital, is a common cognitive deficit found in people with schizophrenia ^34^.

We also found that people with no Foundation Skills were interested in learning more about half of the tasks they could not perform. Given that lack of interest is a commonly reported barrier to digital engagement ^9^, this suggests that people with SMI might have the motivation to learn new skills if appropriate support becomes available. Future studies should seek to consult with those with lived experience to explore motivation in beginning and sustaining digital skills training, preference for training mode and setting, as well as barriers to engaging with web-based resources specifically related to SMI conditions (e.g. fluctuating levels of wellness, low energy and motivation, paranoid thoughts, suspiciousness and concerns about privacy and difficulty processing information; ^35^), in order to develop accessible skills training programs tailored to the needs of this population.

Older people in this study were less likely to have Foundation Skills, as also found in the general population ^23^. This is unsurprising, as age is traditionally associated with less digital engagement in SMI ^14-16^ and general population ^36, 37^ alike. People with psychosis-spectrum disorders were at greater risk for not having Foundation Skills compared to those with bipolar disorder, after adjusting for people’s age or employment status. This supports our previous findings that during the pandemic restrictions people with bipolar disorder were more likely to use the Internet a lot for their daily activities compared to people with psychosis ^16^. Cognitive and occupational disfunctions that are common in schizophrenia ^38^might not facilitate development of digital skills. It might also be that specific SMI-related barriers to digital engagement (e.g. reduced concentration, hallucinations, or paranoid ideas; ^13, 39^) are more common in people with psychosis than bipolar disorder.

The SPIDER study is not free of limitations. We used a non-random sample that might lead to respondent biases and non-representative samples. Skills were self-evaluated by participants rather than completing objective tests. Self-evaluation might be susceptible to other fluctuating factors (e.g. current mood), as well as response and social desirability bias (e.g. n = 134, 53.8% of the sample completed the survey over the phone with a researcher). It should be noted though that organising objective tests, especially in the context of the pandemic restrictions, would present several practical challenges. Participants were also assessed on the basis of pre-determined tasks, rather than personalised tasks related to the specific activities they want/need to complete in their everyday lives. However, that would affect the comparability of results across different populations and existing datasets. Regarding digital access, we did not examine whether the devices that people owned were current enough or had to be shared with other members of the household, as it often happened during the pandemic ^40^. Despite the high rates of device ownership in our sample, real unobstructed access might be undermined by obsolete technologies or multi-usership.

## Conclusion

This study highlighted deficits in digital skills in the SMI population that are worse than in the general population. This might suggest that an already socially disadvantaged population are at risk for further exclusions due to digital skills deficits, as many health services and social connections have moved to digital platforms during 2020 / 2021. Importantly and worryingly this includes vital health services many of which might continue to operate digitally into the future. Services including the NHS need to be aware and have a responsibility to actively and immediately accommodate for those with SMI who would prefer face-to-face rather than online contact, and whose health will likely suffer through the digital divide. Additionally, further funding is needed for research into a widely available, person-centered, accessible training program, co-produced with end users to identify and remove barriers related to lack of digital skills.

## Supporting information

Detailed methods

## Data Availability

Aggregated data can be made available from authors on request.

https://osf.io/yxvp9/?view_only=680cf833b46942a293b8d5350bbdd89b

## Statements

## Acknowledgements

This report is independent research supported by the National Institute for Health Research Yorkshire and Humber Applied Research Collaboration. The views expressed in this publication are those of the author(s) and not necessarily those of the National Institute for Health Research or the Department of Health and Social Care.

We thank the participants in the SPIDER study and NHS mental health staff for their support with this study.

Ethical approval for this study was granted by the Health Research Authority Northwest – Liverpool Central Research Ethics Committee (REC reference 20/NW/0276). Completing and returning the survey was considered implying consent to participate in the study.

