## Supplementary material for "Measuring the digital divide among people with Severe Mental Ill Health using the Essential Digital Skills framework": Detailed methods

### S1: Methodology of the OWLS study leading to SPIDER

#### Design and Procedures

This study reports on results obtained from the Skills and Proficiency In Digital Essential Requirements study (SPIDER; data collected from January 2021 to March 2021) which drew participants from the Optimising Wellbeing in Self-Isolation study (OWLS; data collected from July 2020 to December 2020). OWLS participants were a sub-group of participants from the Closing the Gap (CtG) study (data collected from April 2016 to May 2020).

The CtG study was a large clinical cohort (N = 9,914) comprising adults (aged 18 years or older) with documented diagnosis of schizophrenia or delusional/psychotic illness (ICD 10 F20.X & F22.X or DSM equivalent) or bipolar disorder (ICD F31.X or DSM equivalent). Ethical approval for the CtG study was granted by West Midlands—Edgbaston Research Ethics Committee (REF 15/WM/0444).

OWLS recruited a sub-cohort from CTG, to explore the effects of the COVID-19 pandemic restrictions on people with severe mental ill health. To be eligible for invitation to OWLS, CtG participants had to have provided contact details and consented to be contacted again, as well as been originally recruited from a clinical site that had the capacity to collaborate with the University of York research team in a new research project. Eligible participants were then organised in groups based on age, gender, ethnicity, and care setting (primary or secondary mental health care) to ensure representation across many sociodemographic groups. From each group, researchers selected a purposive sample of participants that had most recently participated in the CtG study (e.g., recruited in the last two years) ensuring that a range of localities was covered. Recent participation to the CtG was considered important to increase response rates (e.g., the team having current and valid contact details, and participants being familiar with the research team). Locality was used to provide geographical diversity, inviting participants from 17 mental health trusts and six Clinical Research Network (CRN) areas in England, including a mix of rural and urban settings.

Participants who expressed an interest in taking part were provided with an information sheet (read over the phone, or send by email, text message, or post). Those consenting to participate were given the option to complete the survey over the phone with a researcher, online, or completing and returning a hard copy survey sent by post. Ethical approval for the OWLS project was granted by the Health Research Authority Northwest – Liverpool Central Research Ethics Committee (REC reference 20/NW/0276).

#### The OWLS sample

Out of 2,932 participants in the CtG study that were eligible to be invited to OWLS, we selected a purposive sub-sample of 1,166 (39.8 %) participants and successfully contacted 688 (59%). The survey was completed by 367 participants (31.5% of the selected sub-sample and 53.3% of those successfully contacted). The final study sample had a mean age of 50.5 (± 15.69) years old and it included 51.0% men, 47.4% women, 1.6% transgender, 17.7% people from other than White ethnic background and 48.5% residing in high/very high deprivation areas in the country. The primary diagnosis was psychosis (51.2%). The survey was completed online by 121 participants (33%) and over the phone or via the post by 246 (67%). In terms of location, 51.2% of participants were recruited from the North of England, 5.7% from East Midlands/Anglia, 10.4% from London, and 32.7% from the South of England.

#### The OWLS survey

Participants in OWLS self-reported information on the following domains: 1. Mental and physical health and wellbeing, 2. Experiences using healthcare services, 3. Health-related behaviours (e.g., smoking, alcohol, diet, physical activity, etc.), 4. COVID-19 specific experiences (e.g., being ill with COVID-19, having to self-isolate, etc.), 5. Loneliness and social support, 6. Use of the Internet and digital devices, and 7. Employment and financial status.

The surveys are available in the OWLS repository in the Open Science Framework at https://mfr.osf.io/render?url=https://osf.io/qpfn6/?direct%26mode=render%26action=download%26mode=render f

**Figure 1: Flow Diagram - OWLS**

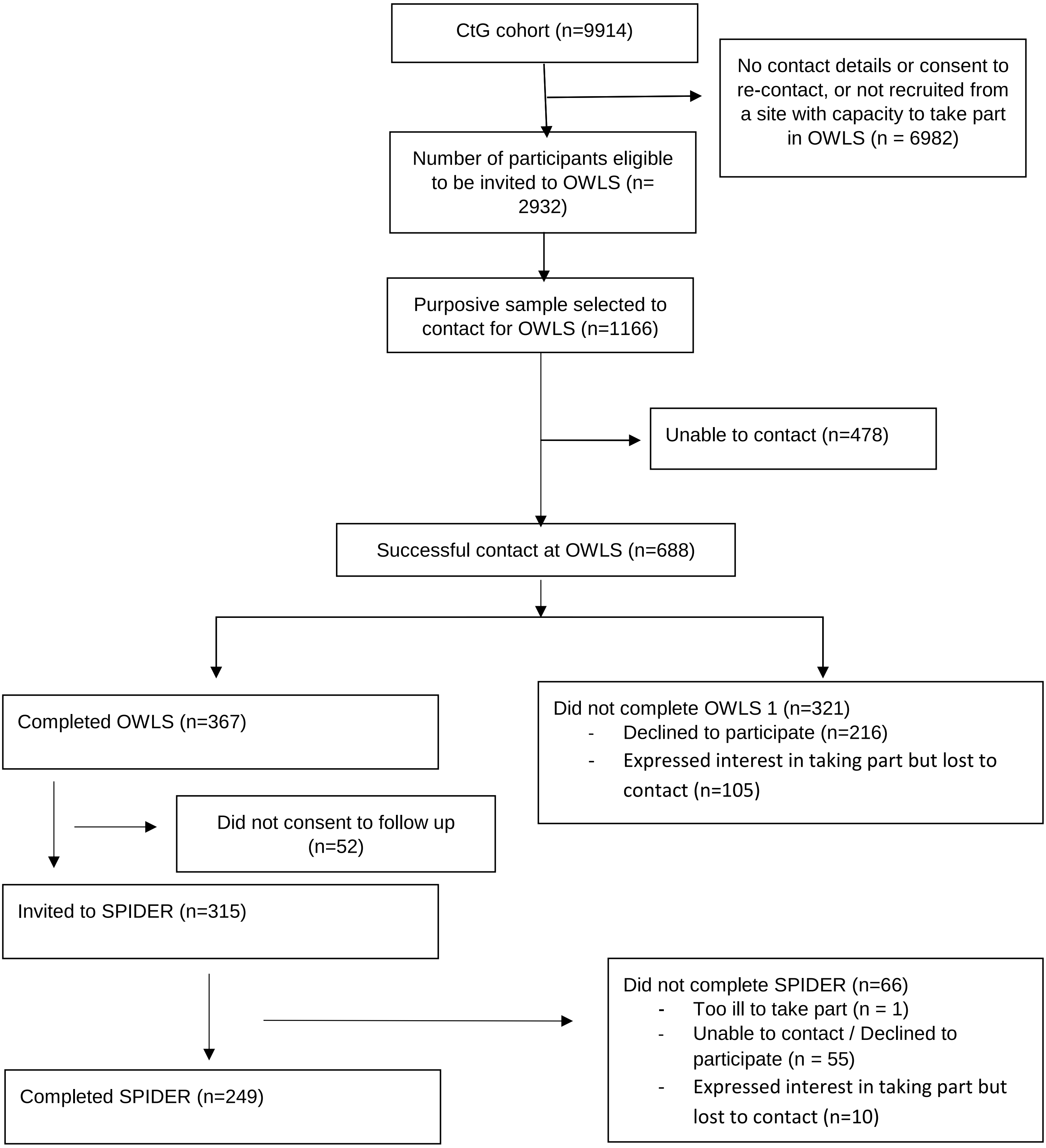
